# EVALUATION OF HEALTH-RELATED QUALITY OF LIFE IN PATIENTS WITH DIABETES IN DIFFERENT CARE SETTINGS A CROSS SECTIONAL STUDY IN ALAIN, UAE

**DOI:** 10.1101/2021.06.19.21259165

**Authors:** Mariam Salem Khamis Al kaabi, Bushra khamis Obaid Al Kaabi, Fatima Mohammed Ahmed Al Marzooqi, Shaima Ghazi Al Murri, Latifa Mohammad Baynouna AlKetbi

**Author notes:** Corresponding author’s address and email ID: Abu Dhabi Healthcare Services, Po Box: 85010, AlAin, 00971564991105.

## Abstract

**Purpose:** This study aims to assess health-related quality of life (HRQOL) in type 2 diabetic patients across four domains—physical, psychological, social, and environmental—and explore the possible determinants of these domains.

**Methods:** Using a cross-sectional study design, 397 type 2 diabetic patients in Alain city were interviewed using validated questionnaires in three different care settings: primary, secondary and private health care facilities. The WHO Quality of Life instrument, generalized anxiety disorder score, and Patient Health Questionnaire (PHQ9) were employed.

**Results:** The participants comprised 270 females (68%) and 127 males (32%), with 68.9% in the 41–65 years age group; 49.6% were married. The most common comorbidities were dyslipidemia (69.3%), hypertension (61%), and osteoarthritis (24.7%). On a scale of 0 to 100, the highest QOL mean score was reported in the social relationship domain (78.3), followed by the environmental (77.7), psychological health (74.2), and physical health (70.7) domains.

The risk of depression was a strong determinant of poor physical health. Social factors have great impact on a patient’s health and well-being. We noticed no difference in HRQOL outcome between primary, secondary, and private health care facilities.

**Conclusion:** The social and mental health domains were the most influential in HRQOL among the participants. This finding supports targeting QOL assessments of patients with type 2 diabetes at the regular chronic diseases clinics and in the planning of population health management programs to ensure the best outcomes.

**Plain English Summary:** Diabetes mellitus is considered a major cause of morbidity and mortality worldwide. In our study we are aiming to explore difference in quality of life and quality of care in patients who access different health care facilities, primary health care clinics versus patients following in secondary care center and private center in Alain. By doing so, we will expand the knowledge regarding health related quality of life of people with diabetes in the UAE and opens the door for future international collaborative research in Diabetes.

## Introduction

The burden of diseases cannot be assessed with mortality alone; both morbidity and mortality are important indicators as well. Studies have used mortality statistics and a variety of estimates of morbidity to measure the state of the health of populations, including hospital admissions, outpatient attendance, and prevalence estimates ^[1]^. The contribution of the disease itself to the outcome is only partial, while other influential factors can increase or decrease the total burden of any disease. Diabetes mellitus is considered a major cause of morbidity and mortality worldwide. Its complications affect multiple organ systems and can affect the life of the patients across multiple domains—physical, psychological, social, and environmental. The World Health Organization (WHO) projected that 300 million people will suffer from diabetes mellitus by 2025 ^[2]^. Figures from the International Diabetes Federation (IDF) in February 2020 revealed that 15.4% of the UAE adult population have type 2 diabetes with a total number of 1.2 million cases ^[3]^. To mitigate the impact of this condition on population health, it is essential to assess the burden of diabetes in the UAE through a well-developed tool.

Though frequently used or relied on, clinical indicators can be insufficient to capture the overall well-being of an individual with diabetes mellitus ^[4–6]^. The psychosocial burden of living with diabetes mellitus can significantly affect self-care behaviors, leading to poor glycemic control, long-term complications, and poor quality of life (QOL) ^[1]^. Thus, a measurement of quality of life would better represent disease burden, level of care, and patient’s overall satisfaction.

Health-related quality of life (HRQOL) refers to the physical, psychological, social, and environmental domains of health ^[7]^. As type 2 diabetes is a chronic disease with complications affecting multiple organ systems, it can have a significant impact on patient’s quality of life and psychological health. Quality of life is an important outcome of medical treatment and has been characterized as “the ultimate goal of all health interventions” ^[8]^.

Assessing quality of life in patients with diabetes via regular implementation of the WHO HRQOL questionnaire can guide us to enhance our patient care tailored to the affected domain, resulting in a reduction of disease burden, associated costs, morbidity, and mortality [9].

A better understanding of HRQOL would help in health care performance assessment and resource allocation ^[1,10,11]^. Moreover it can facilitate planning of future services and management of diabetes at the regional, national, and international levels ^[12]^. It helps not only to assess a patient’s status and QOL, but also to identify patients at high risk of poor health outcome and QOL. The aim of this study is to evaluated the HRQOL and associated socio-demographic and clinical variables among patients with type 2 diabetes.

## Methods

### Design and setting

This study adopted an observational cross sectional design and was conducted on 397 patients with type 2 Diabetes mellitus, across the city of Alain, UAE. Three healthcare settings were selected for comparison of HRQOL, primary, secondary and private healthcare facilities. Preexisting psychiatric condition or low socioeconomic status are possible confounders.

### Questionnaires used

The questionnaires used in the study were the WHO Quality of life (WHOQOL), Generalized anxiety disorder (specifically, GAD7) score and Patient Health Questionnaire (PHQ9).

The WHO Quality of life (WHOQOL) questionnaire is used to interview patients with type 2 diabetes mellitus to assess their HRQOL. In our study, we used the WHOQOL-BREF questionnaire, which is a shorter version of the WHOQOL-100 survey tool developed by the WHO to evaluate aspects of HRQOL. WHOQOL was developed for multicultural comparisons of QOL and has been translated into more than 40 languages. It has been used in several countries worldwide, including the United States of America, Netherlands, Poland, Bangladesh, Thailand, India, Australia, Japan, Croatia, and Zimbabwe ^[13]^ It assesses four QOL domains: physical health, psychological, social, and environmental. The WHOQOL-BREF contains a total of 26 questions, and the answers are scored on a five-point Likert scale. Each domain’s total score is subsequently transformed to a 0–100 scale, where 100 denotes the highest, and 0, the lowest HRQOL. The mean score of items within each domain is used to calculate the domain score. Mean scores are then multiplied by 4 in order to make domain scores comparable with the scores used in the WHOQOL-100 tool ^[13]^. Permission was taken from the developer of the WHOQOL-BREF questionnaire, which was subsequently translated and validated in the Arabic language ^[13]^.

The disadvantage of WHOQOL is that it is administered from an individual point of view and not from the viewpoint of health care professionals. The WHOQOL instruments cover areas that are not discussed in medical interventions, such as personal relationships and social activities ^[14]^.

The WHOQOL questionnaire was translated and validated in the Arabic language following permission received from the developer. GAD7 and PHQ9 have already been translated and used in the Ambulatory Healthcare Services (AHS) health care centers; these are allowed to be used freely by their developer.

### Data collection and analysis

Data collection was done where patient attended their follow up clinic waiting area either primary, secondary or private healthcare facility. Consent was taken with the questionnaire and was signed by the patient, through direct interviews conducted by the four researchers. The preliminary demographic data on gender, age, nationality, marital status, smoking status, level of education, number of visits to diabetes clinics, medications (oral, insulin, or both), administration of medication (by self, family member, housekeeper), and comorbidities (hypertension, dyslipidemia, osteoarthritis, anxiety, depression) were collected at the start of the interview. The PHQ9, GAD7, and HbA1C scores were identified from the patient chart. The PHQ9 and GAD7 items were added to the interview if it was not available.

The inclusion criteria are as follows: individuals with type 2 diabetes mellitus; in the age group of 18 years and above; attending one of the three health care settings selected for the study; competent in Arabic or English; both UAE nationals and non-nationals. The exclusion criteria are as follows: patients with type 1 diabetes mellitus or other illnesses not related to diabetes mellitus, such as cancer; new history of traumatic events, such as death of loved ones; and newly divorced.

Data were analyzed using the Statistical Package for Social Sciences software or SPSS version 15 (SPSS Inc., Chicago, IL, USA). Data were described using means, standard deviations, and proportions wherever appropriate. Differences in means between controlled (HBA1C <8%) and not-controlled (HBA1C ≥ 8%) diabetes were analyzed using *t*-test. Multiple binary logistic regressions were used to determine factors associated with poor glycemic control.

The study was approved by the Research Ethics Committee in Alain, UAE.

## Results

This study included a total of 397 (127 males [32%] and 270 females [68%]) patients with type 2 diabetes mellitus. Their age ranged from 18 years and above. About half of the patients (49.6%) were in the age group of 41–65 years. The majority of the patients were UAE nationals (78.9%) and more than two-thirds were married (68.9%). Table 1 presents the participants’ socio-demographic, clinical, and other relevant characteristics. The total number of patients who were on insulin across the three health care settings (primary, secondary, and private) was 47 (11.9%). While 295 patients (74.5%) were on oral hypoglycemic medication, and 53 patients (13.4%) were on both insulin and hypoglycemic agents. The most common comorbidities were dyslipidemia (140; 69.3%), followed by Hypertension (110; 61.5%), and osteoarthritis (98; 24.7%). The majority of the participants, including both males and females, were non-smokers (356; 89.9%). Regarding the control of diabetes, the study categorized participants with HbA1c levels less than 8 (75% of the study sample) as having controlled diabetes, and those with HbA1c of 8 or more (remaining 23.8%) as having uncontrolled diabetes.

**Table 1.** Demographic Characteristics of the study participants.

|  |  | CENTER |  |  | Total |
| --- | --- | --- | --- | --- | --- |
|  |  | Primary | Secondary | Private |  |
| GENDER | Female | 143(36%) | 102(25.7%) | 25(6.3%) | 270(68%) |
|  | Male | 60(15.1%) | 57(14.4%) | 10(2.5%) | 127(32%) |
| AGE | 20-40 | 36(9.1%) | 21(5.3%) | 2(0.5%) | 59(14.9%) |
|  | 41-65 | 113(28.5%) | 63(15.9%) | 21(5.3%) | 197(49.6%) |
|  | >65 | 54(13.6%) | 75(18.9%) | 12(3.0%) | 141(35.5%) |
| NATIONALITIES | UAE | 152(38.7%) | 129(32.8%) | 29(7.4%) | 310(78.9%) |
|  | Non-UAE | 51(13%) | 26(6.6%) | 6(1.5%) | 83(21.1%) |
| STATUS | Single | 7(1.8%) | 12(3%) | 0(0%) | 19(4.8%) |
|  | Married | 154(38.9%) | 95(24%) | 24(6.1) | 273(68.9%) |
|  | Widow | 35(8.8%) | 46(11.6%) | 9(2.3%) | 90(22.7%) |
|  | Divorced | 6(1.5%) | 6(1.5%) | 2(0.5%) | 14(3.5%) |
| EDUCATION | Illiterate | 59(14.9%) | 63(15.9%) | 13(3.3%) | 135(34%) |
|  | Primary | 53(13.4%) | 29(7.3%) | 8(2%) | 90(22.7%) |
|  | Preparatory | 27(6.8%) | 19(4.8%) | 4(1%) | 50(12.6%) |
|  | High School | 38(9.6%) | 20(5%) | 6(1.5%) | 64(16.1%) |
|  | University | 26(6.5%) | 28(7.1%) | 4(1%) | 58(14.6%) |
| SMOKER | YES | 19(4.8%) | 16(4%) | 5(1.3%) | 40(10.1%) |
|  | NO | 184(46.5%) | 142(35.9%) | 30(7.6%) | 356(89.9%) |

**Table 1. Demographic Characteristics of the study participants (continued)**
|  |  | CENTER |  |  | Total |
| --- | --- | --- | --- | --- | --- |
|  |  | Primary | Secondary | Private |  |
| VISIT | First | 54(14.1%) | 49(12.8%) | 3(0.8%) | 106(27.7%) |
|  | 2-4 | 26(6.8%) | 17(4.5%) | 1(0.3%) | 44(11.5%) |
|  | >4 | 109(28.5%) | 92(24.1%) | 31(8.1%) | 232(60.7%) |
| MEDICATION | Insulin | 22(5.6%) | 25(6.3%) | 0(0%) | 47(11.9%) |
|  | Oral hypoglycemic medication | 168(42.4%) | 98(24.7%) | 29(7.3%) | 295(74.5%) |
|  | None | 1(0.3%) | 0(0.0%) | 0(0.0%) | 1(0.3%) |
|  | Both | 12(3.0%) | 35(8.8%) | 6(1.5%) | 53(13.4%) |
| CARE OF MEDICATION | Self | 152(38.3%) | 100(25.2%) | 31(7.8%) | 283(71.3%) |
|  | Family member | 36(9.1%) | 38(9.6%) | 2(0.5%) | 76(19.1%) |
|  | Made | 15(3.8%) | 21(5.3%) | 2(0.5%) | 38(9.6%) |
| HYPERTENSION | Yes | 110(27.7%) | 112(28.2%) | 22(5.5%) | 244(61.5%) |
|  | No | 93(23.4%) | 47(11.8%) | 13(3.3%) | 153(38.5%) |
| DYSLIPIDEMIA | Yes | 140(35.3%) | 106(26.7%) | 29(7.3%) | 275(69.3%) |
|  | No | 63(15.9%) | 53(13.4%) | 6(1.5%) | 122(30.7%) |
| OSTEOPOROSIS | Yes | 28(7.1%) | 26(6.6%) | 13(3.3) | 67(17.1%) |
|  | No | 173(44.1%) | 130(33.2%) | 22(5.6%) | 325(82.9%) |
| OSTEOARTHRITIS | Yes | 46(11.6%) | 36(9.1%) | 16(4%) | 98(24.7%) |
|  | No | 157(39.6%) | 122(30.8%) | 19(4.8%) | 298(75.3%) |
| ANXIETY RISK | Mild | 192(49.0%) | 149(38%) | 34(8.7%) | 375(95.7%) |
|  | Moderate | 7(1.8%) | 3(0.8%) | 1(0.3%) | 11(2.8%) |
|  | Moderately Sever | 3(0.8%) | 3(0.8%) | 0(0.0%) | 6(1.5%) |
|  | Sever | 2(0.5%) | 0(0.0%) | 0(0.0%) | 2(0.5%) |
| DEPRESSION RISK | Normal | 189(48.3%) | 139(35.5%) | 32(8.2%) | 360(92.1%) |
|  | Mild | 6(1.5%) | 12(3.1%) | 3(0.8%) | 21(5.4%) |
|  | Moderate | 4(1%) | 3(0.8%) | 0(0.0%) | 7(1.8%) |
|  | Moderately Sever | 1(0.3%) | 0(0.0%) | 0(0.0%) | 1(0.3%) |
|  | Sever | 2(0.5%) | 0(0.0%) | 0(0.0%) | 2(0.5%) |

Among the four domains in the WHOQOL-BREF instrument, on a scale of 0 to 100, the highest QOL mean score was reported in the social relationship domain (78.3 SD±16.0), followed by the environmental (77.7 SD±15.5), psychological health (74.2 SD17.6) and physical health (70.7 SD±20.4) domains (Table 2).

**Table 2.** Health related quality of life (HRQOL) domain scores.

| Table 2. Health related quality of life (HRQOL) domain scores |  |  |  |  |  |
| --- | --- | --- | --- | --- | --- |
|  | N | Minimum | Maximum | Mean | Std.<br>Deviation |
| Physical Health | 397 | 6.00 | 100.00 | 70.7229 | 20.41770 |
| Psychological Health | 395 | 6.00 | 100.00 | 74.2380 | 17.64337 |
| Social Health | 339 | 0.00 | 100.00 | 78.2566 | 16.03442 |
| Environment | 395 | 31.00 | 100.00 | 77.6734 | 15.48184 |

Seeking care at any of the primary, secondary, and private health care facilities was not found to be determinant of the HRQOL outcome. This was evident from the regression analysis of the determinants of the four QOL domains reported in Table 3. The overall composite of the better quality of life, average of the four domains, was only significantly associated with the following factors: male gender (B 4.371, p<0.003), higher education (B 1.036, p<0.046) UAE national (B-3.284, p<0.035) non-smoker status (B 5.982, p<0.008), lower HbA1C (B -0.967, p<0.024), younger age (B -4.019, p<0.001), self-dependence in taking medication (B 2.215, p<0.026), and lower depression risk (B -1.819, p<0.001).

**Table 3.** Regression analysis of the determinants of the four QOL domains.

|  |  |  |  |  |
| --- | --- | --- | --- | --- |
| Model | Unstandardized Coefficients |  | Standardized Coefficients | Sig. |
|  | B | Std. Error |  |  |
| Dependent Variable: Psychological Health |  |  |  |  |
| Environment | 0.666 | 0.046 | 0.619 | <0.001 |
| Physical Health | 0.234 | 0.035 | 0.287 | <0.001 |
| PHQ9 | -1.115 | 0.18 | -0.185 | <0.001 |
| AGE | 2.794 | 0.745 | 0.135 | <0.001 |
| Employment | -2.553 | 1.064 | -0.084 | 0.017 |
| Insulin | -3.491 | 1.387 | -0.073 |  |
| Dependent Variable: Physical Health |  |  |  |  |
| Psychological Health | 0.515 | 0.077 | 0.420 | <0.001 |
| Social Health | 0.220 | 0.058 | 0.208 | <0.001 |
| Osteoarthritis | 4.314 | 1.470 | 0.108 | 0.004 |
| Environment | 0.193 | 0.095 | 0.147 | 0.042 |
| Care of medication | -2.420 | 1.190 | -0.082 | 0.043 |
| Dependent Variable: Environment |  |  |  |  |
| Psychological Health | 0.513 | 0.043 | 0.586 | <0.001 |
| Physical Health | 0.180 | 0.037 | 0.241 | <0.001 |
| Age | -2.978 | 0.767 | -0.132 | <0.001 |
| Nationality | -2.445 | 1.025 | -0.067 | 0.018 |
| Employment | 2.265 | 1.101 | 0.068 | 0.040 |
| Visit frequency | 1.176 | 0.485 | 0.070 | 0.016 |
| GAD7 | -0.466 | 0.214 | -0.067 | 0.030 |
| Dependent Variable: Social Health |  |  |  |  |
| Environment | 0.658 | 0.059 | 0.529 | <0.001 |
| PHQ9 | -1.987 | 0.282 | -0.287 | <0.001 |
| Visit frequency | 3.133 | 0.602 | 0.179 | <0.001 |
| Physical Health | 0.195 | 0.046 | 0.207 | <0.001 |
| GAD7 | 1.224 | 0.406 | 0.121 | 0.003 |
| Employment | -2.967 | 1.213 | -0.085 | 0.015 |
| **Table 3. Regression analysis of the determinants of the four QOL domains continued** |  |  |  |  |
| Dependent Variable: Composite |  |  |  |  |
| PHQ9 | -1.819 | 0.275 | -0.319 | <0.001 |
| AGE | -4.019 | 1.208 | -0.204 | 0.001 |
| Care of medication | -4.142 | 1.228 | -0.180 | 0.001 |
| Gender | 4.371 | 1.464 | 0.159 | 0.003 |
| Smoker | 5.982 | 2.233 | 0.138 | 0.008 |
| HBA1C | -0.967 | 0.427 | -0.116 | 0.024 |
| Nationalities | -3.284 | 1.546 | -0.101 | 0.035 |
| Education | 1.036 | 0.518 | 0.120 | 0.046 |

The important determinants of each domain are shown in Table 3. For the psychological domain, the significant determinants include age, as a higher domain score was associated with increasing age (B2.79, p<0.001). Unemployment work status was significantly associated with a lower psychological domain score (B-2.55, p<0.017). Furthermore, the psychological domain score was found to be significantly associated with higher environmental and physical domain scores. Depression risk is another determinant of the psychological domain as a lower risk of depression (B -1.026 p<0.001) was found in patients with better psychological QOL. Insulin use was associated with lower psychological health (B-3.49, p<0.001).

Improvements in the physical health domain score were associated with a lower dependence on others in the administration of medication (B -2.42, p<0.043) or having osteoarthritis (B 4.31, p<0.004). Psychological health (B 0.515, p<0.001), environmental (B 0.193, p<0.001) and social health domain scores (B 0.220 p<0.001) were better among patients with better physical health.

Among the determinants of the environmental domain, age seems to be of great significance since a higher environmental domain score was associated with a lower age (B -2.98, p<0.001).

For a 1% increase in the social domain score, there was a significant 1.99% (p<0.001) reduction in PHQ9 depression risk score as well as an increase in the scores of the physical domain by 0.195% (p<0.001) and environmental domain by 0.658% (p<0.001). Furthermore, better social QOL was associated with higher visit frequency to the center (B 3.133, p<0.001), better physical health (B 0.195, p<0.001), and higher risk of anxiety (B 1.224, p<0.001).

While investigating the relationship between psychological QOL domain and the PHQ9 and GAD scores, this study aims to evaluate whether including the QOL questionnaire in patient evaluation better predicts depression. The findings suggest that the comparison between psychological QOL domain and PHQ9 significantly correlated. Figure 1 depicts that a higher psychological domain score often reflects a lower risk of depression by PHQ9.

**Fig. 1.**
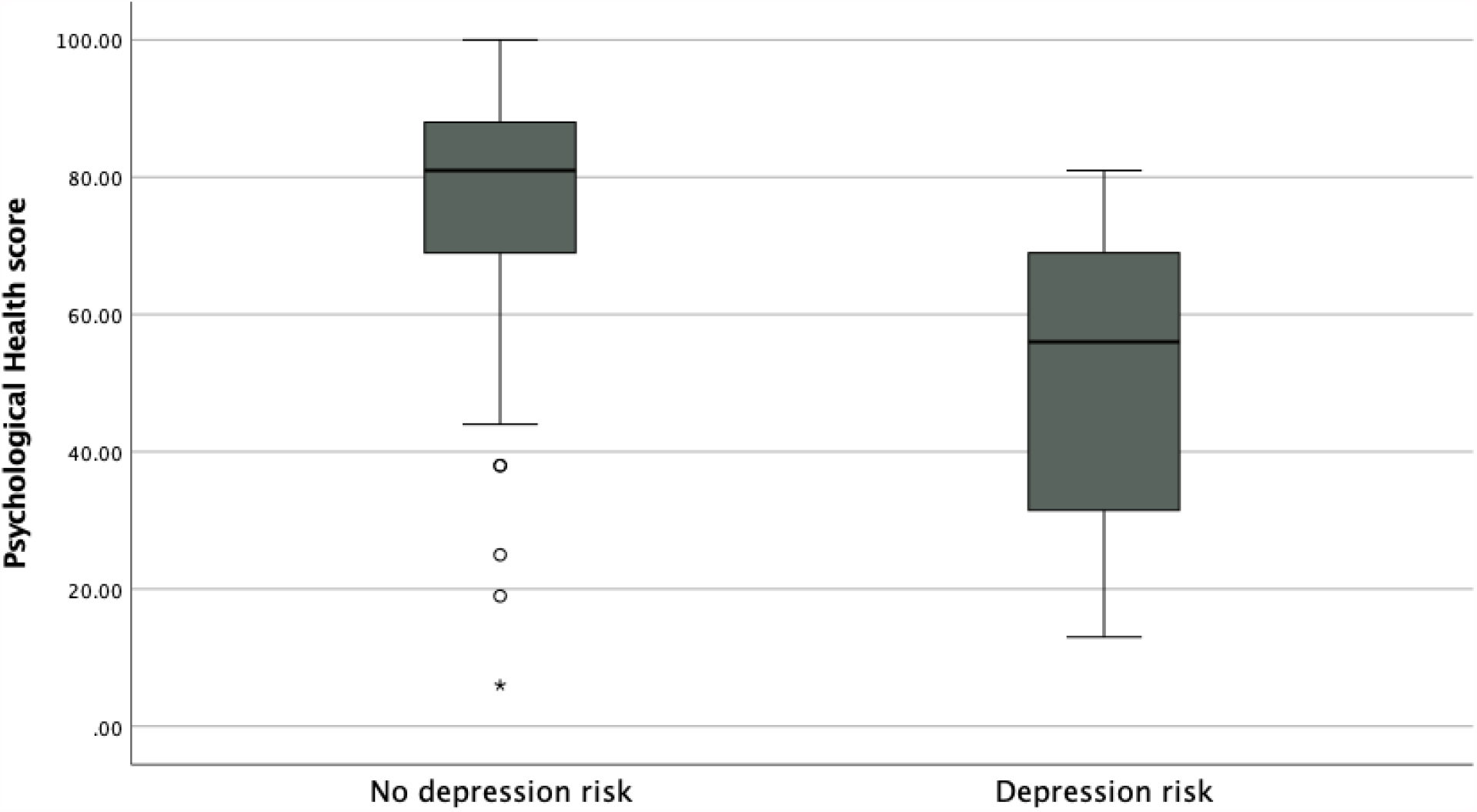
Psychological domain score and by PHQ9 depression score

Figure 2 illustrates that with an increasing risk of depression, the scores for three domains (psychological, social, and environmental) decreased, but the physical domain score remained consistent throughout.

**Fig. 2.**
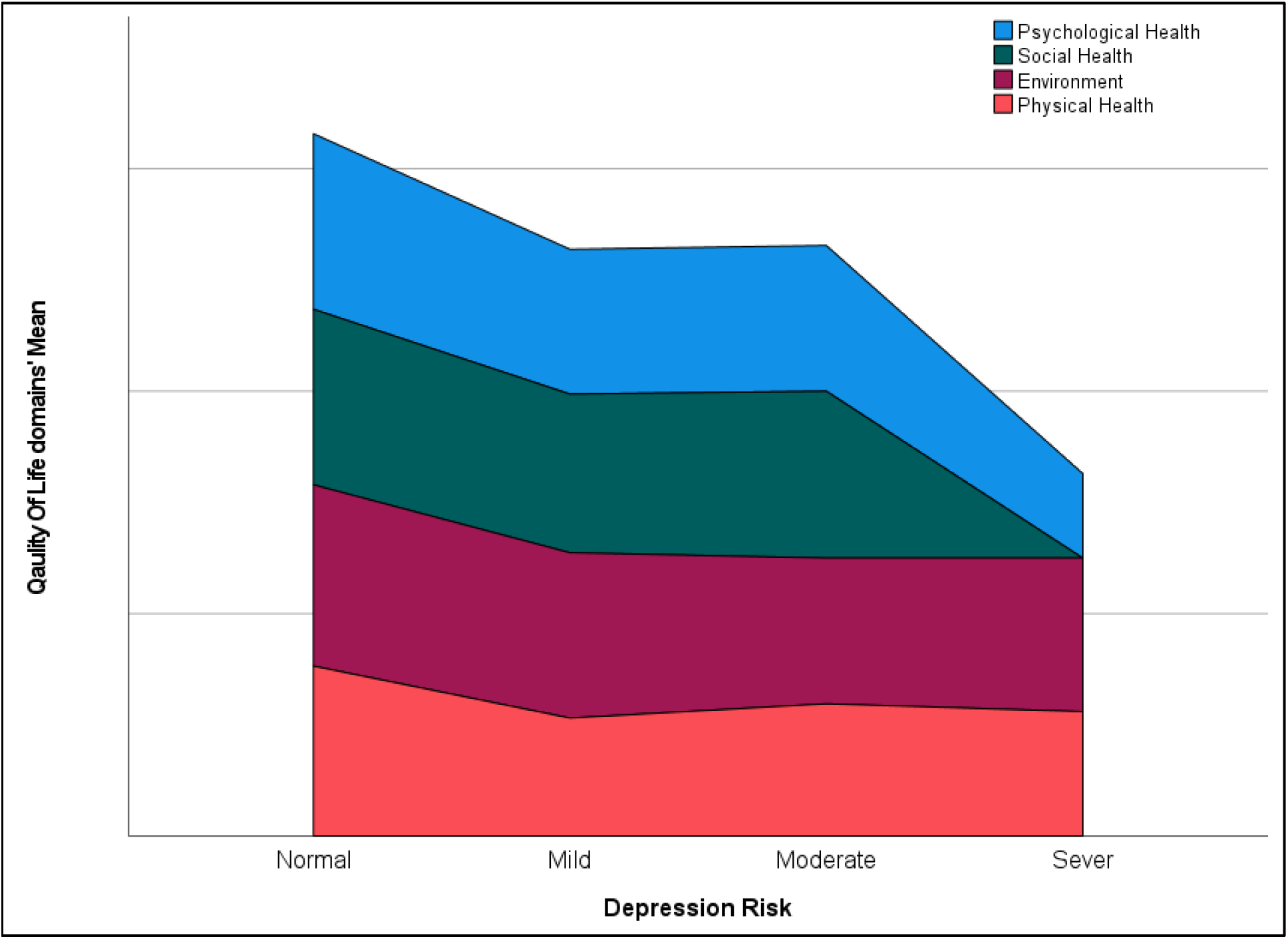
QOL domains with increasing risk of depression

The environmental score was found to be higher among younger patients (B-2.978, p<0.001), UAE nationals (B-2.445, p 0.018), those who are employed (B 2.265, p 0.04), those with higher visit frequency (B 1.176, p 0.016), and lower GAD7 scores (B -0.466, p 0.03).

The social health domain score was higher among patients who reported lower depression risk (B-1.987, p<0.001), but have higher risk of anxiety (B 1.224, p 0.003). Moreover, it was higher among those with higher number of visits to the health care centers (B 3.133, p<0.001) and those who are unemployed (B-2.967, p 0.015).

Overall, the composite HRQOL score was positively associated with lower risk of depression, younger patients, being male, non-smokers, lower HbA1C, UAE nationals, and higher education.

Figure 3 shows that a better control of DM (HbA1c<8) is associated with a higher environmental domain score. Similarly, it was found that seeking care at a primary, secondary, or private facility is not a determinant of better diabetic control.

**Fig. 3.**
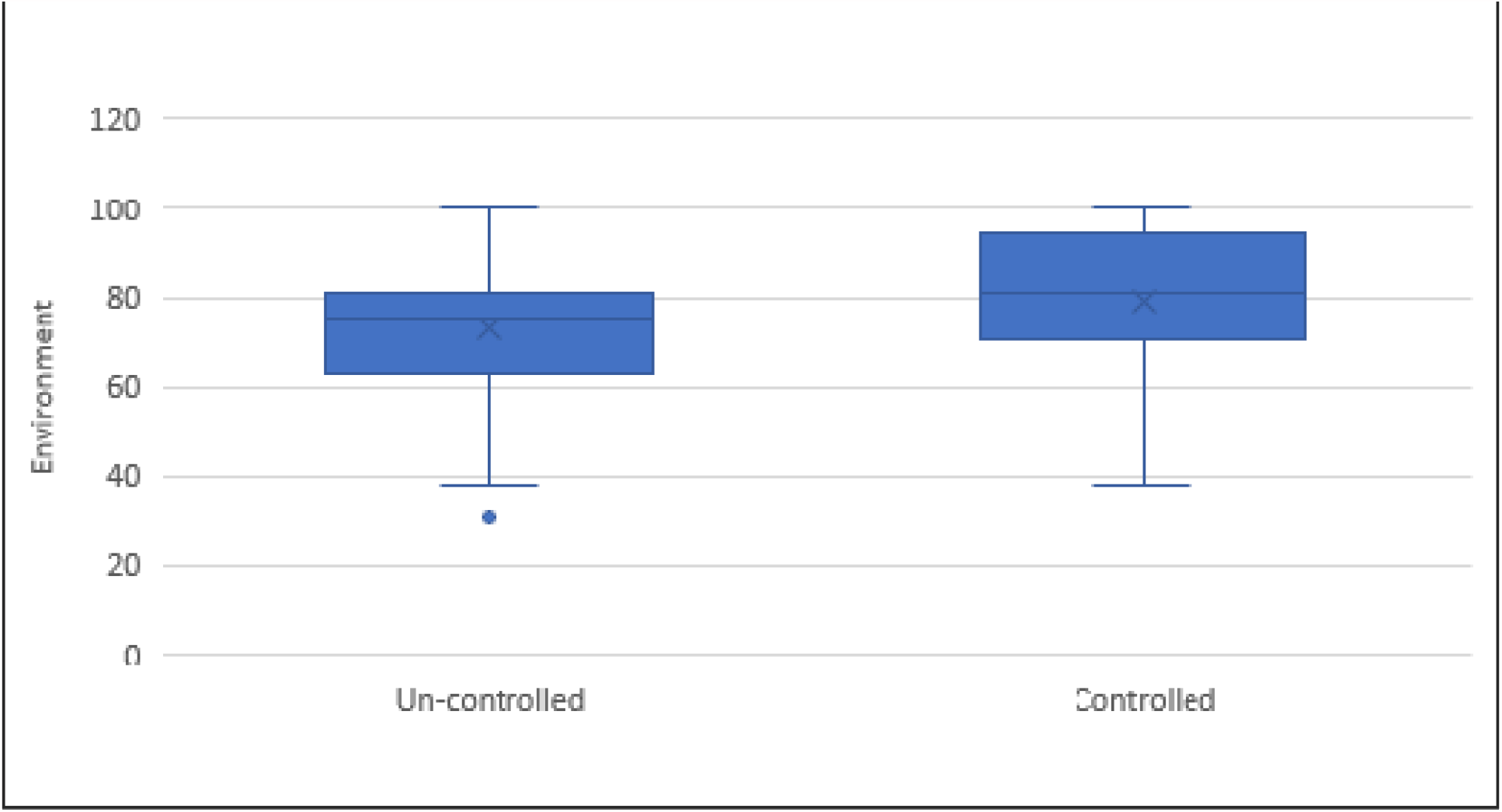
Box plots; control of DM and environmental domain score

## Discussion

This study revealed that of the four WHOQOL-BREF domains, the least affected QOL domain is the social domain and the most affected is the physical health domain. This implies that the participants were relatively more satisfied with their social support system.

In this population, age was associated with a higher HRQOL, in terms of the composite and all individual domain scores, except that of the social domain. This is similar to a 2020 study, conducted in Saudi Arabia, which showed that there was a negative association between age and HRQOL among patients with diabetes ^[15]^. Their results showed 30% of older patients with type 2 diabetes had severe impairment in the HRQOl, while the proportion was 16% among patients aged less than 50 years.

In Germany, a 2014 study showed that socioeconomic status is positively associated with better HRQOL among patients with a chronic disease. 40% of patients who have a primary education or lower education reported severe impairment in HRQOL compared to patients with a higher educational qualification ^[16]^. Similarly, the current study revealed that lower education was associated with a worsening composite HRQOL.

Studies from the US show that improved HbA1c is associated with better HRQOL, similar to the findings of the current study^[17]^. Given the strong evidence of the benefit of diabetes control, the finding that a better QOL is associated with lower HbA1C only necessitates interventions targeting better control among patients with diabetes.

Nevertheless, it may not be that straightforward. Depression is one of the stronger predictors of medical outcomes than the physiological and metabolic measures (such as the presence of complications, body–mass index, and HbA1c) ^[18]^. It is possible that there is a bidirectional association between depression and diabetes as an increase in depression is associated with an increased risk of diabetes complications, such as microvascular and macrovascular complications, which can further result in mental health impairment and reduced HRQOL^[19]^. Also, current evidence supports that psychosocial issues are critical to good diabetes care ^[18]^. Targeting the modifiable factors that influence the four domains, such as depression, anxiety, and frequency of visits, may affect quality of life and subsequently, the patient may become more empowered to self-manage and make the right choices. Choices such as adherence and lifestyle changes were not included in this study, but can be investigated in future research. It is worth noting that hospital-based care is more expensive than primary health care ^[20]^.

This study revealed that there is no difference in QOL between primary care and specialty care in diabetes. This finding underscores the role of primary care in the care of patients with diabetes, further highlighting the cost-effectiveness of primary care ^[21]^.

Furthermore, insulin may improve QOL by reducing hyperglycemia symptoms and micro vascular complications. However, insulin may also negatively affect QOL through side effects, such as hypoglycemia and weight gain, psychological distress, stigmatization, and the limitation of social activities.

Many studies have examined the effect of insulin use on QOL in patients with diabetes; a 2019 study in Austria reported no statistical difference in the WHOQOL-BREF score between patients using insulin or patients treated with oral therapy^[22]^This result was like that of other studies which used the WHOQOL-BREF instrument ^[23,24]^Nevertheless, other studies have concluded that insulin therapy improves QOL [18,25]. In our study, insulin use was associated with lower psychological health.

Another important finding is that maintaining good physical health and, specifically, good management of osteoarthritis might contribute to a better independence in medication use (as observed in our population).

Good social life showed a significant positive influence in reducing the risk of depression and having a better quality of life. Thus, a focus on enhancing the social determinants of health— including socioeconomic status, education, neighborhood and physical environment, employment, and social support networks, as well as access to health care—will aid in improving HRQOL of patients with type 2 diabetes. Assessing and targeting social determinants of health were found to be essential for management of diabetes ^[26]^.

As noted in the results, the social health domain score was higher among those with a higher visit frequency to the health care centers and those who are unemployed. This finding could be explained by the amount of free time available for social interaction and leisure activities in the unemployed group, for example due to retirement. The factors to consider for the employed group include sedentary type of work and long working hours. Long working hours might be associated with adverse health outcomes ^[27]^.

Moreover, different occupational types differ in physical and mental requirements, which might affect health status of the person—for example, employees that work in different time shifts such as health care workers. A focus on work–life balance will lead to improved mental well-being and QOL ^[28]^. Additionally, QOL was found to be a predictor of mortality ^[29]^.

A study conducted in Gaborone, Botswana observed that females and older adults had a poorer QOL score, similar to the findings of this study ^[8]^. A possible reason could be the increasing age-related health issues such as osteoarthritis. As for gender differences, other factors such as hormonal status (menopause) and psychosocial factors that were not measured in this study might have a negative impact on some HRQOL domain scores. This is an important avenue for future research to better guide care planning ^[30]^.

This necessitates increased efforts to explore the factors that determine the lower HRQOL in women, and consequently, more targeted approaches can be implemented to improve HRQOL in women ^[30]^.

A limitation of this study includes the risk of reporting bias in the questionnaire. Further, some participants found it difficult to immediately understand the response categories for certain items in the questionnaire; most of the time, the questions had to be explained more clearly, especially for older adults.

Another limitation in our study is the lack of comparison to a healthy population. A study in the International Journal of Caring Sciences reported that the quality of life in patients with diabetes mellitus was lower compared to normal healthy individuals, and that diabetes had a negative outcome on HRQOL. However, in our study, we did not investigate the relationship between the duration of the disease and QOL in patients with diabetes as that can be a determinant as well. Assessing the duration of the disease may help us better understand how severely the HRQoL will be affected. However, another study examined the duration of the disease, compared to HRQOL, and showed that the longer the duration of the disease, the higher will be the risk of developing complications, thereby reducing QOL ^[1]^.

## Conclusion

In our population, social support and mental health are of paramount importance for improving QOL of patients with diabetes. We recommend that this study be conducted on a wider scale with a larger sample size to better represent our population. Implementing WHOQOL questionnaires as a part of regular chronic disease clinic care and in the planning of population health management programs can enhance patient care and ensure best outcomes.

## Data Availability

All data referred to in the manuscript of this study can be made available from the corresponding authors upon request.

## Abbreviation

(HRQOL): Health-related quality of life
(PHQ9),): Patient Health Questionnaire
(specifically, GAD7): Generalized anxiety disorder
(WHO): The World Health Organization
(AHS): Ambulatory Healthcare Services
(IDF): International Diabetes Federation.

## Ethical Approval and consent to participate

The study was approved by AHS and Tawam Ethics committee

## Consent for publication

All investigators consented to publish **Availability of supporting data** Data available when requested

## Author’s contributions

Conceptualizations: All authors. Data collected BAK, MAK, FAM, SH

Data analysis LBK, writing Manuscript

